# Genome-Wide Insights into the Genes and Pathways Shaping Human Foveal Development

**DOI:** 10.1101/2025.06.18.25329836

**Authors:** Callum Hunt, Ha-Jun Yoon, Alvin Lirio, Kayesha Coley, Jun Wang, Nick Shrine, Jianming Shao, Gail DE Maconachie, Zhanhan Tu, Jonathan H Zippin, Pirro G Hysi, Christopher J Hammond, Ala Moshiri, Rui Chen, Martin D Tobin, Chiara Batini, Mervyn G Thomas

## Abstract

Here we report the first genome-wide association study of foveal pit depth. In a cohort of 61,269 individuals, we identified 123 genome-wide significant loci associated with pit depth, including 47 novel associations not previously linked to macular traits. Using 12 complementary variant-to-gene mapping strategies, we prioritised 128 putative causal genes, 64 of which have not previously been implicated in foveal development. Our findings reveal previously unrecognised biological influences on foveal morphogenesis, including retinoic acid metabolism (implicating *CYP26A1* for the first time in human foveal development), extracellular matrix and cytoskeletal dynamics, and retinal cell fate determination. In addition, rare-variant analysis uncovered two further gene associations, including *ESYT3*, a gene not previously linked to foveal structure. Together, these results provide new insights into the genetic architecture and molecular pathways underlying human foveal development, and offer a foundation for future functional studies aimed at characterising foveal development and disease.

## Introduction

High-acuity vision in humans relies on the proper development of the fovea, a highly specialised retinal region that is morphologically distinct from the peripheral retina. The foveal architecture is defined by the excavation of a distinctive pit, the displacement of inner retinal layers, the formation of an avascular zone, and the dense packing of specialised cone photoreceptors within a rod-free central region (Thomas et al., 2022). These structural and functional differences demand a unique cellular composition and gene expression profile, distinguishing the fovea not only in its morphology but also in its underlying molecular characteristics (Yan et al., 2020).

Disruptions in foveal development cause foveal hypoplasia (FH), which is most commonly observed in individuals with albinism and is often accompanied by other ocular abnormalities, including misrouting of the optic chiasm and nystagmus (Kuht et al., 2022). The known spectrum of genes associated with FH are primarily involved in various aspects of retinal pigmentation, including melanosome formation and maintenance, melanin synthesis, and related signalling pathways. However, the molecular mechanisms through which pigmentation defines proper foveal development remain poorly understood (Bakker et al., 2022).

The fovea can be visualised using optical coherence tomography (OCT), a powerful imaging tool which facilitates high-resolution and layer-wise observation of the macula and fovea (Huang et al., 1991; Kondo et al., 2018). OCT is the primary tool that is used to diagnose FH clinically and enables the severity of FH to be determined based on the presence or absence of foveal architecture such as the foveal pit (Thomas et al., 2011). Abnormal foveal morphology, as visualised by OCT, is also observed in acquired conditions such as epiretinal membranes, vitreomacular traction, macular degeneration, diabetic maculopathy, inflammatory disease and macular holes.

Aside from this clinical utility, OCT scans have also been used to derive quantitative measures such as retinal layer thickness, which have proven to be powerful traits in genome-wide association studies (GWAS) (Gao et al., 2019; Currant et al., 2021; Currant et al., 2023; Zekavat et al., 2024; Sergouniotis et al., 2024; Jackson et al., 2025). Collectively, these studies underscore the value of image derived traits in identifying genetic signals relevant to macular health while also highlighting the retina as a potential indicator of systemic health. However, the direct quantification of foveal pit morphology in both normal and abnormal fovea for similar analyses remains unexplored.

In this study, we measured foveal pit depth in the UK Biobank (UKB) cohort and conducted the first GWAS of this trait. To translate genetic findings into biological insights, we applied a comprehensive variant-to-gene approach leveraging 12 lines of evidence, including regulatory annotations, functional effects, and rare-variant associations. We extended our rare variant analysis to the exome-wide level to uncover additional genes implicated in foveal development. In doing so, we aimed to gain mechanistic insight into the pathways shaping foveal development and disease.

## Methods

### UK Biobank data and optical coherence tomography (OCT)

Phenotypic and genetic data available from UK Biobank (UKB) were accessed and analysed as part of this research, under UKB application 85881. UKB has approval from the North West Multi-centre Research Ethics Committee (MREC) as a Research Tissue Bank approval (REC reference: 21/NW/0157).

The UKB is a large longitudinal cohort study which recruited approximately 500,000 people aged between 40-69 years across the United Kingdom (Bycroft et al., 2018). In addition to collecting baseline participant characteristics, a subset of participants underwent ophthalmic assessments including OCT examination. OCT scans in UK Biobank were obtained using the TOPCON 3D OCT1000 Mark 2 instrument which constructs a three-dimensional (3D) scan of the retina in a 6 mm^2^ area, by combining 128 cross-sectional B-scan images per eye across the macula (Keane et al., 2016).

### Deriving foveal pit depth in UKB participants

We analysed the right eye OCT B-scan in UKB participants, selecting the scan that passed cross-sectionally through the foveal centre. The central B-scan from each volumetric OCT dataset was identified using the central frame index provided by the Topcon Advanced Boundary Segmentation (TABS) software (Yang et al., 2010), which corresponds to the scan closest to the anatomical foveal centre. Retinal layer segmentation was performed using TABS (Yang et al., 2010), a proprietary algorithm that automatically segments volumetric OCT data into nine distinct retinal layers. To standardise anatomical alignment across B-scan images, we used the retinal pigment epithelium (RPE), a consistently hyper-reflective layer in OCT imaging, as a reference for B-scan flattening. We trained a custom deep neural network using DeepLabCut (version 2.3.9) with a ResNet-50 backbone to detect eight anatomical landmarks in foveal OCT B-scans. These included two points placed at the peaks of the foveal contour and one point at the pit along the internal limiting membrane (ILM), posterior extent of the inner retinal layers (IRL), and single points on the external limiting membrane (ELM), inner segment ellipsoid (ISe), retinal pigment epithelium (RPE), and Bruch’s membrane/choroid (BM/Choroid) interface. The model was trained using *imgaug* data augmentation pipeline, incorporating histogram equalisation, contrast-limited adaptive histogram equalisation (CLAHE), random rotations (±25°), scale jittering (0.5–1.25×), and embossing to improve robustness to variability in image quality and contrast.

The initial training dataset consisted of 250 high-quality, manually annotated foveal OCT central B-scans from UKB. A separate test set of 50 images was used to evaluate model performance. The trained model was then used to predict landmark locations on an additional set of 2,500 images. Poor-quality B-scans were excluded, and predicted landmarks were refined and manually corrected to create an expanded training set. A final independent test set of 500 images was held out for evaluation.

Training was performed for 500,000 iterations with a batch size of 1, using a staged learning rate schedule: 0.005 for the first 10,000 iterations, 0.02 until 430,000 iterations, followed by 0.002 until 500,000 iterations. The training loss converged to 0.0016, indicating stable convergence and strong model performance (Supplementary Figure 1).

Model predictions were evaluated on the test set using the mean Euclidean distance (in pixels) between predicted and ground-truth landmark positions. Scoremaps (confidence maps) were generated for each landmark, providing a probabilistic spatial representation of the model’s confidence in its predictions. An example scoremap and prediction overlay are shown in supplementary figure 1.

Three of the eight annotated landmarks were placed along the internal limiting membrane (ILM) and were specifically used for foveal pit depth estimation. These included two points at the peaks of the foveal contour and one at the central pit. Foveal depth was defined as the difference between the y-coordinate of the pit and the average y-coordinate of the two peak points. To ensure reliability, predictions were excluded if the average confidence (likelihood) of these three landmarks was below 0.99.

### Genome wide association study (GWAS) of foveal pit depth

Using Phase 3 of the 1000 Genomes Project (1KGP) (Auton et al., 2015), we applied a K-means clustering approach (Shrine et al., 2019) to identify a subset of individuals of European (1KGP-EUR-like) ancestry with quantified foveal pit depth. We then performed association testing with this 1KGP-EUR-like cohort using REGENIE V3.4.1 (Mbatchou et al., 2021) with an additive genetic model, adjusting for age, height, sex, and the first 20 genetic principal components.

For Step 1 of REGENIE, we used directly genotyped array data, retaining variants with <10% missingness, minor allele frequency (MAF) > 0.01, minor allele count (MAC) > 20, and Hardy-Weinberg equilibrium (HWE) *p*-value > 1 × 10□□. For Step 2, we used imputed data generated using a high-coverage (30×) imputation reference panel from Genomics England (GEL) (UKB Data-Field 21008). Association testing was performed on variants with INFO > 0.3 and MAF ≥ 0.01, and for variants with INFO > 0.8 and MAF < 0.01.

To evaluate inflation in test statistics, we calculated the genomic control lambda (λ_GC_) and to assess polygenicity we used the linkage disequilibrium (LD) score intercept using LD Score Regression (LDSC) v1.0.1 (Bulik-Sullivan et al., 2015). Additionally, LDSC was used to estimate the SNP-based heritability (h²) of foveal pit depth using pre-computed LD scores (1000 Genome EUR).

### Identifying foveal pit depth sentinel variants

We identified regions of associated SNPs centred around SNPs showing the most significant association and separated by more than 2MB. For each of these regions we computed 95% credible sets using Sum of Single Effects (SuSiE) regression integrated within Polyfun V1.0 (Weissbrod et al., 2020). LD matrices for Polyfun were produced for each region of interest using LDstore2 (Benner et al., 2017). Where credible sets could not be defined using Polyfun-SuSiE we performed conditional analysis for each region using the cojo-slct function within GCTA-COJO V1.94 (Yang et al., 2012) and produced 95% credible sets for each independently associated variant using the Wakefield method (Wakefield., 2009), setting the prior W to 0.04. Sentinel variants were obtained from each credible set, defined as the variant within the credible set with the highest individual posterior inclusion probability (PIP).

### Variant lookup analysis

We searched PubMed (https://pubmed.ncbi.nlm.nih.gov) and the GWAS Catalog (https://www.ebi.ac.uk/gwas/) to identify previously reported GWAS signals (*P* < 5 × 10□□) associated with the macular region (Gao et al., 2019; Currant et al., 2021, Currant et al., 2023; Zekavat et al., 2024; Sergouniotis et al., 2025; Jackson et al., 2025). We then assessed the extent of linkage disequilibrium (LD) between sentinel variants and published GWAS signals, considering a sentinel variant as previously reported if it had r² > 0.1 with a known signal in UKB. LD calculations were performed using PLINK 1.9 (Purcell et al., 2007; https://www.cog-genomics.org/plink/1.9/). If a previously reported sentinel variant was not present in the GEL imputed dataset, LD with the sentinel variants identified in this study was manually assessed using a European reference population with the LDlink tool. (https://ldlink.nih.gov/?tab=home).

### Variant-to-gene mapping

Sentinel variants were mapped to relevant genes to investigate their function and gain insights into the foveal pit biology. We did so by utilising 12 variant-to-gene criteria and prioritising genes supported by at least two of these criteria. The 12 lines of evidence that we considered are:

#### i. Nearest coding gene to a sentinel variant

The nearest coding gene to each sentinel variant was identified based on the distance (± 1MB) of the sentinel position to the transcriptional start site (TSS) of neighbouring genes. The TSS for neighbouring genes was obtained using the biomaRt package (Drost and Paszkowski., 2017) querying ensemble (Version 112) for the TSS of the canonical transcript.

#### ii. Genic annotation

We determined whether a sentinel variant was located within the exon/promoter/untranslated regions (UTRs) of nearby genes (± 500KB) using the ChIPseeker R package (Yu et al., 2015) according to the gene annotation from the TxDb.Hsapiens.UCSC.hg38.knownGene R library and considering ±1000bp region of gene TSS as the promoter region.

#### iii. In-silico prediction of pathogenicity

To determine if a sentinel variant is a likely pathogenic missense variant within a gene, we queried the variant using the variant effect predictor (VEP) tool (https://www.ensembl.org/info/docs/tools/vep/index.html) and determined that the sentinel variant was annotated as missense and likely to be pathogenic if the CADD PHRED score was ≥ 20.

#### iv/v. Mapping of sentinel variants to cis-regulatory elements

To determine whether a variant is in a *cis*-regulatory region, we identified the potential *cis*-regulatory regions and their target genes utilising a multiomics dataset derived from developing human retinal tissue (Zuo et al., 2024) and adult retinal pigment epithelium (unpublished). We first identified genome-wide open chromatin regions (OCRs) and the regions with differential chromatin accessibility (DARs) across major cell types during retinal development based on snATAC-seq data using the ArchR (Granja et al., 2021) R package (for DAR identification: the function getMarkerFeatures(), getMarkers(), cutOff = “FDR <= 0.05 & Log2FC >= 0.5”). Then we identified OCR-gene pairs through correlating chromatin accessibility of OCRs in the ± 250KB surrounding gene TSS with gene expression, as well as the chromatin accessibility of the promoter respectively with the ArchR R package (getPeak2GeneLinks() for correlating gene expression with chromatin accessibility of surrounding OCRs, corCutOff = 0.5, FDRCutOff = 0.01, resolution = 1. getCoAccessibility() for correlating promoter accessibility with chromatin accessibility of the surrounding OCRs, corCutOff = 0.5, resolution = 1). The union set of OCR-gene pairs identified through the two methods was used to annotate the potential *cis*-regulatory regions in the genome.

#### vi) Polygenic priority score (POPS)

We calculated polygenic priority scores (PoPS) for approximately 18,000 coding genes by combining foveal pit depth GWAS summary statistics with pre-computed gene features and using the EUR superpopulation in the 1000 Genomes Project (Phase 3) as the LD reference panel (Auton et al., 2015). A total of 57,543 features were used in total – 40,546 derived from gene expression datasets, 8,718 based on protein–protein interactions, and 8,479 from biological pathways (Weeks et al., 2023). We prioritised a gene to each sentinel by selecting the gene within ± 250KB which had the highest PoPS score.

#### vii) Overlap with retinal eQTL

To identify whether any foveal pit depth sentinel variants have been previously reported as retinal eQTLs, we performed a direct lookup in published bulk retinal eQTL data (Ratnapriya et al., 2019), restricting our search to variants significant at P < 5×10□□.

#### viii) Overlap with blood pQTL

To determine whether any foveal pit depth sentinel variants have been previously reported as blood pQTLs, we performed a direct lookup using two published pQTL datasets. Specifically, we queried UK Biobank Olink proteomics data (Sun et al., 2023) and deCODE Genetics proteomics data (Ferkingstad et al., 2021). The UK Biobank dataset includes measurements of 2,923 proteins, while the deCODE dataset reports pQTLs for 4,719 proteins using 4,907 aptamers. For UKB Olink we set the significance threshold for protein level associations as *P* < 5 × 10^−8^, but we used a threshold of *P* < 1.8 × 10^−9^ for DeCODE Genetics as reported in the original publication (Ferkingstad et al., 2021). Sentinel variants were then looked-up in each pQTL data set to identify *cis*-pQTLs only, meaning that the TSS of the pQTL gene was within ± 1MB of the sentinel position

#### ix) Nearby mouse knockout orthologs with vision related phenotype

We obtained all human orthologs of mouse knockout genes with a known eye, vision or pigmentation related phenotype within ± 500KB our sentinel variants. Mouse knockout genes were obtained by searching the mouse genome informatics database (https://www.informatics.jax.org/) for phenotype term MP:0005391 and MP:0001186.

#### x) Nearby mendelian disease genes

We identified a list of relevant Mendelian diseases using Orphanet (https://www.orpha.net/) by searching the title and clinical signs of all diseases for the following search terms “fovea, macula, retin, albinism, pigment, photoreceptor, cone, Hermansky-Pudlak, Idiopathic infantile nystagmus” and then obtained a list of all causative genes for the identified disorders. We selected mendelian disease genes among this list, which are within ± 500KB of any sentinel variant.

#### xi/xii Nearby genes implicated by rare variant analysis

We identified genes nearby to sentinels (± 500KB) that are implicated by rare-variant associations in foveal pit depth using whole-exome sequencing data. We considered single rare-variant associations and gene-based collapsing analysis as independent predictors (Duffy et al., 2024). We performed both rare-variant association testing and collapsing analyses using REGENIE V3.4.1 (Mbatchou et al., 2021), excluding variants which are determined by UKB as having poor sequencing coverage (https://dnanexus.gitbook.io/uk-biobank-rap/science-corner/whole-exome-sequencing-oqfe-protocol/generation-and-utilization-of-quality-control-set-90pct10dp-on-oqfe-data) and using age, sex, height and the first 20 genetic principal components as covariates.

For single rare-variant association testing we restricted analysis to rare exonic variants (MAF < 0.01 and MAC >=3) and individuals of 1KGP-EUR-like ancestry. We prioritised genes which were implicated by a rare variant association meeting a suggestive significance threshold of *P* < 5 × 10^−6^. Rare variant collapsing analysis was performed using a previously defined set of variant annotation masks (Backman et al., 2021), and using four allele frequency bins (0.01,0.001,0.0001,0.00001). A single *p*-value for each gene analysed was obtained using the *p*-value omnibus test GENE_P within REGENIE and significance was assessed at a Bonferroni adjusted threshold of *P* < 2.86 × 10^−6^, accounting for the 17,471 protein coding genes tested.

### Assignment of putative causal genes to functional groups

We investigated the likely role of putative causal genes in foveal development using GeneCards (https://www.genecards.org) and published literature in PubMed (https://pubmed.ncbi.nlm.nih.gov). Where possible genes were assigned to a functional group based on their involvement in 1) pigmentation or the RPE, 2) metabolism, 3) photoreceptors, 4) retinal cell fate, 5) retinal vessel development, or 6) the cytoskeleton and extracellular matrix (ECM).

### Enrichment analysis of putative causal genes with Metascape

We used the gene annotation tool Metascape (Zhou et al., 2019) to investigate the biological processes enriched among the putative causal genes identified in our GWAS study. Specifically, we utilised Metascape to identify the top 20 curated biological pathways based on Gene Ontology (GO) enrichment and pathway databases, namely KEGG Pathway, Reactome Gene Sets, Canonical Pathways, CORUM, WikiPathways, and PANTHER Pathway. Additionally, we assessed the enrichment of genes regulated by key transcription factors using the TRRUST database and examined disease enrichment of the putative causal genes using DisGeNET. In all cases, enrichment was considered significant at a false discovery rate (FDR) threshold of <5%.

### Genetic correlation analysis

Using summary statistics from published genome-wide association studies in European individuals (Supplementary Table 24), we calculated genetic correlations between each trait and foveal pit depth using LD Score regression (LDSC v1.0.1; Bulik-Sullivan et al., 2015) with pre-computed LD scores for European populations. We specifically focused on ocular and pigmentation traits relevant to foveal development and morphology, to assess their shared genetic architecture with foveal pit depth.

### Cross-ancestry association analysis

We performed association testing of the foveal pit depth sentinel variants in two non-EUR populations, African (1KGP-AFR-like, *n* = 1,819) and South Asian (1KGP-SAS-like, *n* = 2,134). Ancestry assignment and association testing followed the methodology previously described for the 1KGP-EUR-like foveal pit depth cohort. We assessed the concordance of sentinel variant effect sizes between the 1KGP-EUR-like cohort and both the 1KGP-AFR-like and 1KGP-SAS-like cohorts, by calculating Pearson’s correlation coefficients.

In addition to association testing with individual foveal pit depth sentinel variants, we evaluated the aggregate genetic contribution to foveal pit depth in both non-EUR ancestry groups using polygenic scores (PGS). These PGS were calculated using the HRC*+*UK10K imputed genetic data (UKB Data-Field 22828) with the PRS-CS v1.1.0 tool (Ge et al., 2019). PRS-CS is a Bayesian method that estimates posterior SNP effect sizes from GWAS summary statistics while automatically learning hyperparameters from the training data, eliminating the need for a validation set. Summary statistics from the 1KGP-EUR-like foveal pit depth cohort served as the training dataset and PGS association analyses were then performed in the 1KGP-AFR-like and 1KGP-SAS-like cohorts using linear regression, adjusting for age, sex and the first 20 genetic principal components.

### Rare-variant discovery analysis

In addition to the rare variant analysis used to support variant-to-gene analysis, we also sought to identify genes implicated by rare variant associations that were not implicated by the GWAS. For single rare-variant analysis, we applied a stringent genome-wide significance threshold of *P* < 5 × 10□□, but we retained the Bonferroni-adjusted significance threshold (*P* < 2.86 × 10^−6^) for collapsing analysis.

## Results

### GWAS of foveal pit depth

We used deep learning to quantify foveal pit depth in UK Biobank participants and identified 61,269 individuals of 1KGP-EUR-like ancestry for association testing – the first large-scale genetic study of this specific foveal trait. Foveal pit depth ranged from 2.7μm to 231.1μm, with a median foveal pit depth of 116.4μm. We conducted a GWAS of foveal pit depth with this cohort, testing 35,800,107 variants while adjusting for age, sex, height, and the first 20 genetic principal components. We observed slight genomic inflation (Figure 1), but LD score regression estimated an intercept of 1.033 (standard error [S.E.] = 0.0098), consistent with a polygenic architecture rather than confounding. The SNP-based heritability (h²) was estimated at 0.2858 (S.E. = 0.0302), indicating that common genetic variation explains nearly 29% of the variance in foveal pit depth demonstrating a substantial polygenic contribution to foveal morphology for the first time.

**Figure 1.**
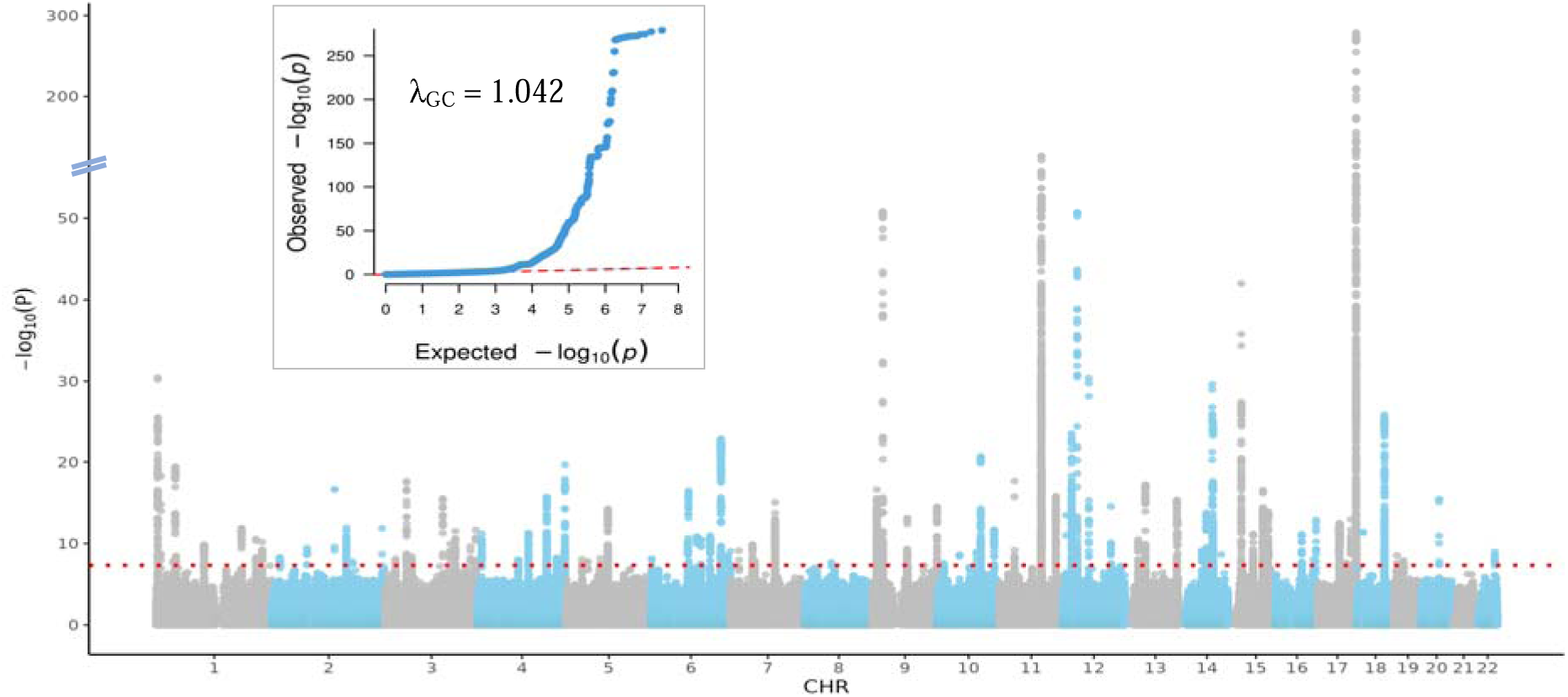
Manhattan and quantile-quantile (QQ) plots for genome-wide association analysis of foveal pit depth. The Manhattan plot displays genome-wide association results, with each point representing a genetic variant. The x-axis indicates chromosomal position, and the y-axis shows the −log□□(p-value). The red dotted lines represent the genome-wide significance threshold (*P* < 5× 10□^8^). The QQ plot compares observed versus expected –log□□(p-values) under the null hypothesis, with the genomic inflation factor (λ_GC_) provided as a measure of population stratification or confounding.

Through fine-mapping analysis we identified 123 independently associated sentinel variants (Supplementary Table 2), and by assessing their linkage disequilibrium (LD) with previously reported signals for macular traits, determined that 47 represent novel associations with the macular region (Supplementary Table 3). Fourteen of the 123 sentinel variants had a posterior inclusion probability (PIP) > 95%, and 48 had a PIP ≥ 50%. The median number of variants per 95% credible set was 8 (Supplementary Table 1).

### Identifying 128 putative causal genes influencing foveal pit depth

We implicated 128 putative causal genes through variant-to-gene analysis as contributors to foveal pit depth variation (Figure 2). To achieve this, we leveraged 12 independent lines of variant-to-gene evidence. Our approach integrated sentinel-variant annotation and rare-variant association evidence, explored the potential regulatory role of sentinel variants on gene expression using QTL data and single-cell transcriptomics data, and utilised functional evidence from Mendelian disease and mouse knockout databases (Supplementary Tables 4– 16).

**Figure 2:**
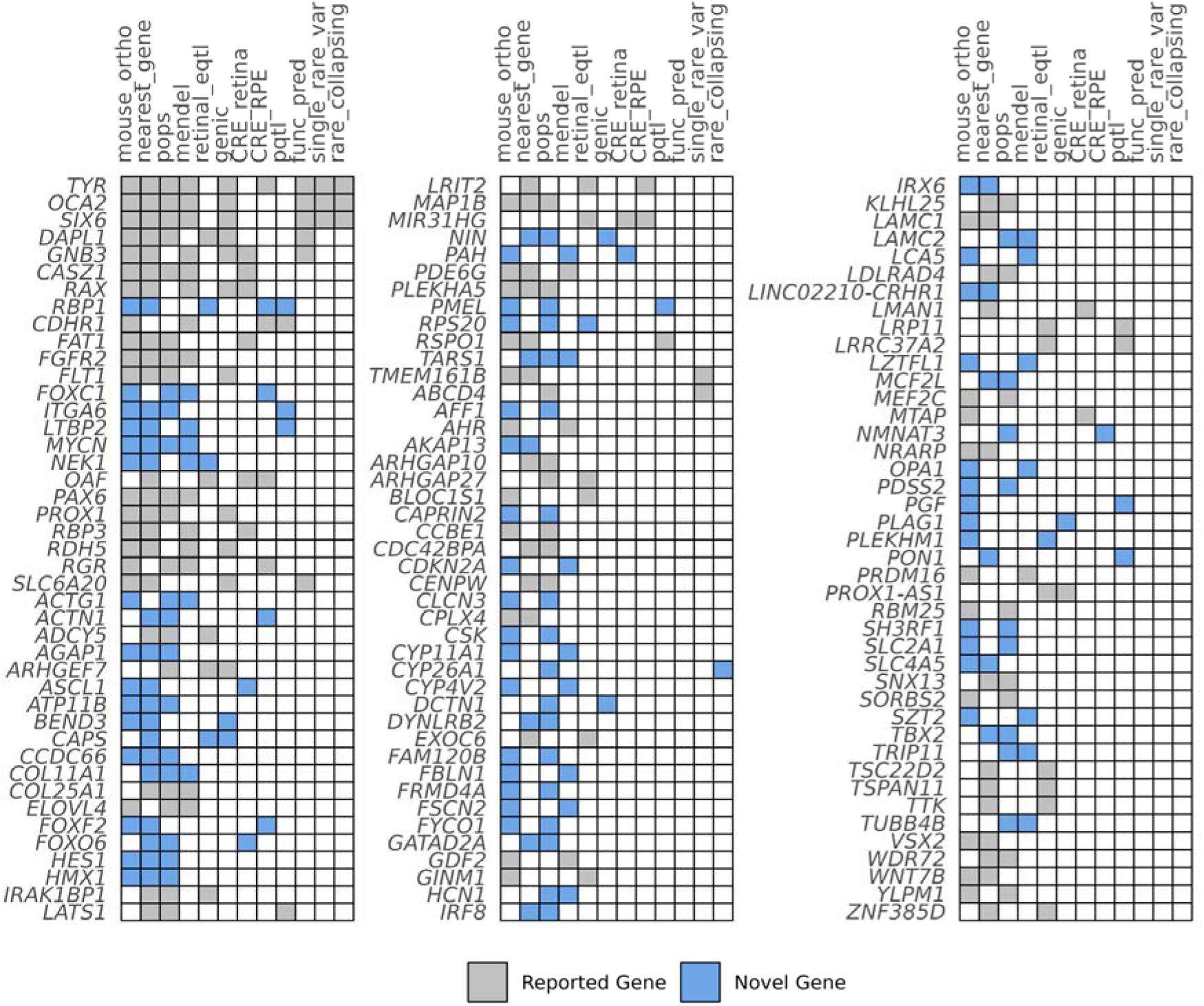
Summary of the variant-to-gene evidence supporting putative causal genes. Evidence for novel gene associations with foveal pit depth is highlighted in blue, while genes previously reported in GWAS of the macular region or associated with known foveal diseases are shaded in grey. Columns are ordered by the number of genes implicated by each line of evidence.

Cross-referencing the 128 putative causal genes with known FH mendelian disease genes, and the genes prioritised by GWAS reports of the macular region (Gao et al., 2019; Currant et al., 2021; Currant et al., 2023; Zekavat et al., 2024; Sergouniotis et al., 2025; Jackson et al., 2025), revealed that 64 genes had not been previously implicated in foveal pit morphology, 60 are previously implicated with broader macular development and four putative genes are known foveal disease genes (*TYR, OCA2, AHR, PAX6*) (Supplementary Table 17).

### Molecular signatures underlying foveal development

We investigated the likely roles of these 128 putative causal genes in foveal development, categorising them into broad functional groups where possible (Supplementary Table 18, Figure 3). Notably, we uncovered genes in biological pathways not previously implicated in human foveal development including retinoic acid/vitamin A metabolism, retinal cell fate determination, cytoskeletal organisation, and extracellular matrix (ECM) dynamics. Among these newly highlighted pathways, we found 11 novel genes in the ECM/cytoskeleton group, 7 in metabolic pathways (e.g. retinoic acid metabolism), and 5 related to retinal cell fate regulators. Even within well-established foveal pathways, we identified 7 novel genes involved with photoreceptor development, 6 involved in the pigmentation/RPE and 2 involved in blood vessel development. Pathway enrichment analysis using Metascape largely supported this functional classification, while also implicating broader biological processes, including cell polarity, morphogenesis, and organelle localisation (Supplementary Tables 19-20).

**Figure 3:**
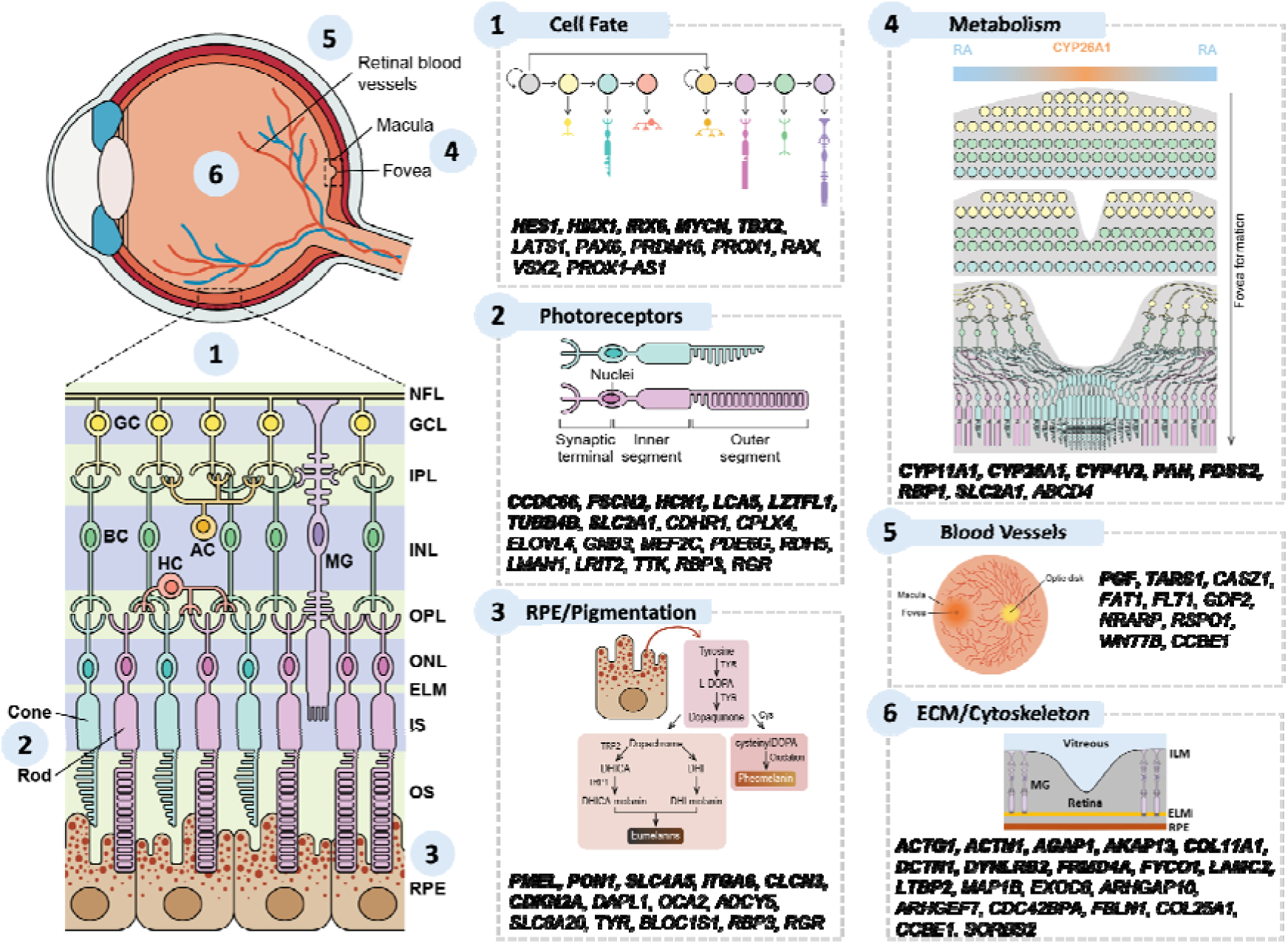
Schematic representation of the retina and foveal region within the human eye. Putative causal genes relevant to prioritised functional groups involved in foveal development are displayed alongside their respective functional categories. Genes highlighted in bold are novel gene associations not reported in previous GWAS of the macular region or as foveal disease genes.

These findings led us to further investigate the regulatory mechanisms that might control the identified pathways. We found enrichment for putative causal genes regulated by two master transcription factors relevant to foveal development (FDR < 5%). *OTX2*, a critical regulator of photoreceptor cell fate (Nishida et al., 2003), regulates *RBP3, PAX6,* and *TYR*. Similarly, *MITF,* a master regulator of melanocyte development (Vachtenheim and Borovanský 2010), governs *PMEL, TYR and OCA2*. Both *PMEL* and *RBP3* represent novel candidate genes implicated in foveal development (Supplementary Tables 17, 21), while *TYR, OCA2* and

*PAX6* are established foveal disease genes.

To further investigate the functional relevance of the putative causal genes, we also performed disease enrichment analysis to identify whether these genes are overrepresented in specific conditions (Supplementary Table 22). Significant enrichment (FDR < 5%) was found for conditions commonly seen alongside foveal hypoplasia such as nystagmus (Kuht et al., 2022), as well as eye disorders affecting broader macular morphology, such as retinitis pigmentosa, glaucoma, age-related macular degeneration, congenital ocular coloboma, retinal dystrophies, and macular dystrophy.

Motivated by these enrichments, we conducted genetic correlation analysis to assess shared genetic architecture between foveal pit depth and select ocular or pigmentation traits (Supplementary Table 24). We observed a weak but statistically significant positive genetic correlation between foveal pit depth and spherical equivalent (*r*_g_ = 0.159, S.E. = 0.025, *P* = 3.17 × 10□^10^). However, no significant genetic correlation was observed with any other traits tested (Supplementary Figure 2).

### Cross-ancestry genetic association with foveal pit depth

Association analysis was performed in two non-European ancestry groups within UKB, 1KGP-AFR-like (*n* = 1,819) and 1KGP-SAS-like (*n* = 2,134), to investigate potential cross-ancestry associations. The median pit depth in these cohorts was 124μm (range: 23μm – 189.5μm) for the 1KGP-AFR-like group and 119.5μm (range: 32.4μm – 188.5μm) for the 1KGP-SAS-like group. We found that the distribution of foveal pit depth differed between the 1KGP-EUR-like cohort and both the 1KGP-AFR-like (W = 43,130,516, P < 8.07 × 10□□¹) and 1KGP-SAS-like (W = 59,754,062, P < 1.37 × 10□¹¹) cohorts, with both populations having larger median foveal pit depth than the 1KGP-EUR-like population. These differences are in line with known variation in foveal pit morphology among people with non-European ancestries (Wagner-Schuman et al., 2011).

We performed association analysis for each foveal pit depth sentinel variant in both non-European ancestry groups (Supplementary Table 23) and evaluated the correlation of effect sizes between the 1KGP-EUR-like ancestral group and two non-European ancestry groups. The strongest correlation was observed between 1KGP-EUR-like and 1KGP-SAS-like ancestries (r = 0.510, P = 3.806 × 10□□), while a weaker correlation was found with 1KGP-AFR-like ancestry (r = 0.403, P = 7.245 × 10□□). We further generated polygenic scores (PGS) for people of 1KGP-SAS-like and 1KGP-AFR-like ancestry using weights derived from the GWAS of the 1KGP-EUR-like population and tested for association of these PGS with foveal pit depth in both non-EUR populations. We found that the PGS were significantly associated with variation in foveal pit depth in both the 1KGP-AFR-like and 1KGP-SAS-like ancestry individuals. This association was highest in the 1KGP-SAS-like population (SD change in phenotype per SD change in PGS (β) = 4.9042; 95% CI: [4.0745, 5.7340]; P = 3.93 ×□10^−30^), and more modest in the 1KGP-AFR-like population (SD change in phenotype per SD change in PGS (β) = 2.6229; 95% CI: [1.7174, 3.5285]; P = 1.59 ×□10^−8^) with a consistent direction of effect. This cross-population association analysis offers the first evidence that the genetic architecture underlying foveal pit depth is, in part, shared across ancestries.

### Rare-variant gene discovery analysis

To further enhance our gene discovery efforts, we sought to identify genes implicated by rare variants that were not implicated by foveal pit depth sentinel variants (Supplementary Table 14). At a stringent significance threshold of *P* < 5 × 10□□, we identified two additional associations from whole-exome sequencing data. The first variant, 11:66560171-T (*P* = 4.96 × 10LJLJ, allele frequency [AF] = 2.53 × 10LJLJ), is a predicted deleterious missense variant in *ACTN3* (CADD PHRED = 23.6). The second variant, 3:138472579-A (*P* = 4.60 × 10LJLJ, AF = 4.21 × 10LJLJ), is a missense variant in *ESYT3*, with a moderate predicted deleterious effect (CADD PHRED = 7.1).

Both variants exhibited large effect sizes, with 3:138472579-A (*ESYT3*) associated with a mean change of −53.00 µm (SE = 9.15) and 11:66560171-T (*ACTN3*) associated with a mean change of −69.46 µm (SE = 11.85). Plotting retinal thickness maps, demonstrated that participants heterozygous for these variants display larger total macular thickness at the foveal centre relative to the average across the 1KGP-EUR-like cohort (Figure 4A). Foveal pit depth was notably reduced in all heterozygotes (Figure 4B) and observation of OCT scans revealed continuation of the inner retinal layers consistent with FH. *ESYT3* is a novel gene implicated in foveal structure, and we provide new evidence implicating *ACTN3* in foveal development, extending its previously reported association with photoreceptor layer thickness through a common variant (Currant et al., 2023).

**Figure 4:**
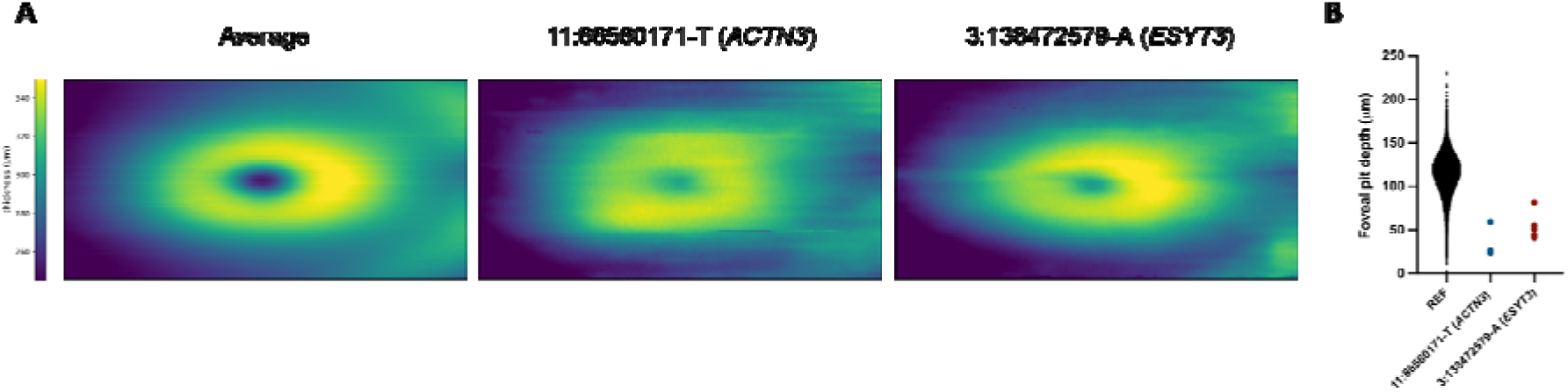
Abnormal foveal morphology in participants heterozygous for rare-variant associations. (A) Average retinal thickness maps across the macula, measured from the internal limiting membrane (ILM) to Bruch’s membrane (BM). Maps are shown for the 1KGP-EUR-like foveal pit depth cohort and for individuals heterozygous for 11:66560171-T (*ACTN3*) or 3:138472579-A (*ESYT3*). (B) Comparison of foveal pit depth between the 1KGP-EUR-like foveal pit depth cohort and participants heterozygous for 11:66560171-T (*ACTN3*) or 3:138472579-A (*ESYT3*).

## Discussion

We conducted the first GWAS of foveal pit depth, identifying 123 common genetic signals of which 47 are novel. These signals mapped to 128 putative causal genes, including 64 which are novel candidates implicated in foveal development. Rare variant analysis identified two additional genes, including *ESYT3*, which has not previously been associated with foveal structure. Exploring the function of prioritised genes revealed previously unrecognised roles for vitamin A metabolism, retinal cell fate determination, and extracellular matrix/cytoskeletal dynamics in human foveal pit morphology, and uncovered novel genes in both these newly discovered and previously described foveal pathways. By investigating both novel and previously reported genetic loci and genes, our results also provide specific evidence supporting the involvement of previously known loci in shaping the fovea, extending their relevance beyond general retinal or macular development (Gao et al., 2019; Currant et al., 2021; Currant et al., 2023; Zekavat et al., 2024; Sergouniotis et al., 2025; Jackson et al., 2025).

To further understand the biological mechanisms underlying these findings, we examined key genes and pathways implicated by our GWAS in greater detail. Among the genes involved in vitamin A metabolism, those involved in retinoic acid (RA) signalling are of particular importance – a connection not previously demonstrated in humans. A lack of RA defines the high-acuity zone in chick (da Silva et al., 2017), and retinal organoid studies demonstrate that an absence of RA defines cone photoreceptor fate at the expense of rods (Sanjuro-Soriano et al., 2022), aligning with the high cone density and rod absence at the foveal centre. *CYP26A1*, which we implicate by rare-variant collapsing analysis, degrades RA, and in mice its combined action with Cyp26c1 helps establish RA gradients in the developing retina (Sakai et al., 2001). Other vitamin A-related genes identified include those involved in the conversion between forms of vitamin A (*RDH5, RGR*) and transport or binding of vitamin A (*RBP1, RBP3*) which may have a regulatory role in this process. Collectively, our findings suggest that vitamin A metabolism, particularly the degradation of RA by *CYP26A1*, may influence human foveal development by shaping the spatial patterning of cone photoreceptors.

Our GWAS also identified genes involved in broader retinal cell fate determination beyond photoreceptors. Notably, we implicate two novel developmental regulators, *HES1* and *HMX1*, which are expressed in retinal progenitor cells and direct the spatio-temporal patterning of the retina (Wall et al., 2009; Boisset and Schorderet., 2012). The identification of genes involved in various aspects of retinal cell fate determination and temporal patterning is underappreciated in foveal development and suggest that the correct proportion and spatial arrangement of many retinal cell types (not just photoreceptors) is a requirement for proper definition of the foveal architecture.

We also implicate genes involved in cytoskeletal and extracellular matrix dynamics as novel determinants of the foveal architecture. This includes a novel gene *ACTN3* that is traditionally linked to muscle performance (Pickering and Kiely., 2017), which we implicated by a rare variant association with a large effect size in heterozygotes. Additional investigation into the role of cytoskeletal and matrix dynamics could shed more light on the mechanical factors that influence the formation of the foveal pit. This could have relevance to other known structural determinates of the foveal development, including the definition of the FAZ, which plays a supportive role in development of the foveal depression (Springer and Hendrickson 2004). We identified novel genes involved in the blood vessel development, reinforcing the importance of FAZ specification in foveal pit development.

As expected, retinal pigmentation emerged as a key determinant of foveal pit depth in our GWAS, with implicated genes involved in various aspects of pigmentation, including pigment synthesis (*TYR*), melanosome development (*PMEL, BLOCS1*), and organelle pH regulation (*OCA2*). The role of common genetic variation in pigmentation genes is increasingly recognised, and notably, certain common hypomorphic alleles of *TYR* have been shown to account for a portion of the missing heritability in individuals with albinism (Loftus et al., 2023). Our findings point to the broader genetic influences of common signals on melanosome biology and pigment synthesis (Kuht et al., 2022). For example, we implicate *PMEL*, a key regulator of melanosome morphology and localisation within the RPE (Burgoyne et al., 2015). Disruptions in melanosome formation are a hallmark of syndromic albinism (Kuht et al., 2022), often driven by mutations affecting the synthesis of lysosome-related organelles, including the BLOC protein complex. In line with this, our GWAS also implicates *BLOC1S1*, identifying it as a novel foveal-development gene within the BLOC complex.

Our analysis also highlights a novel metabolic connection in foveal development, centred on RPE function. While the role of pigmentation in foveal development is still being elucidated, its general importance in the function and maintenance of the RPE, as well as RPE signalling is well described (Bakker et al., 2022). Our GWAS identifies several genes involved in RPE function, including the proline transporter *SLC6A20*, which we identified through a predicted pathogenic variant in this study. Though *SLC6A20* is implicated in previous GWAS of the macular region, it may be particularly important for foveal RPE, as its function could be linked to *SLC38A8*. Mutations in *SLC38A8* cause ocular characteristics of albinism without affecting pigmentation (Poulter et al., 2013) and lead to altered glutamate and glutamine metabolism (Weiner et al., 2025). Notably, proline elevates glutamate and glutamine content in the RPE (Yam et al., 2019), suggesting a potential metabolic interplay between *SLC6A20* and *SLC38A8*. Further exploration of amino acid metabolism, particularly within the retinal pigment epithelium, could provide new insights into foveal development, highlighting a potentially underrecognised role in FH and the broader ocular abnormalities seen in albinism.

Our study, while focused on gene discovery for foveal pit depth, also identifies genes and pathways clinically relevant to Mendelian forms of FH, offering a bridge between common and rare disease genetics. Given the high SNP-based heritability of foveal pit depth (∼30%), our results offer a valuable opportunity to explore how common genetic variation and polygenic architecture may modulate the expressivity of conditions that are traditionally considered monogenic. This raises the hypothesis, now testable using our data, that polygenic background contributes to the phenotypic variability observed in Mendelian foveal diseases (Fahed et al., 2020).

Consistent with this potential for genetic modifiers, several of the putative causal genes identified in our GWAS are also implicated in a wide range of ocular disorders, including conditions commonly co-occurring with FH such as nystagmus (Kuht et al., 2022), as well as retinal degenerations like retinitis pigmentosa, glaucoma, AMD, and macular dystrophy. While this overlap at the gene level suggests biological convergence, genome-wide genetic correlation analysis indicates that the broader polygenic architecture of foveal pit depth is largely distinct from that of common ocular disorders. We observed no significant genome-wide correlation with any tested ocular disease, reinforcing the notion that the genetic programme governing foveal development is, in large part, genetically separable from disease processes. Although the absence of shared heritability with these ocular disorders, many of which are age-related, may also reflect limited prevalence of advanced disease in the GWAS cohorts. A weak but significant genetic correlation with spherical equivalent was observed, which may reflect shared developmental pathways, as FH and reduced pit depth have been associated with microphthalmia, nanophthalmos, and hyperopia: phenotypes linked to disrupted ocular growth (Thomas et al., 2022).

Our findings emphasise the need to develop effective foveal animal models which are currently limited, to better understand and validate the pathways shaping foveal development. It would also be interesting to understand how the important novel mechanisms we implicate in our study, such as retinoic acid metabolism and modifications to retinal cell fate are related, if at all, to variation in retinal pigmentation. This could be relevant to therapeutic strategies in albinism aiming to restore retinal pigmentation and understand the potential consequences for visual function, especially since foveal cone photoreceptors are immature at birth and continue to specialise during the first few years of life (Thomas et al., 2022), providing a therapeutic window to target their maturation.

Nevertheless, we acknowledge several limitations of our study. First, we were unable to replicate our primary GWAS findings in an independent dataset due to the scarcity of comparable biobanks with both genotypic and OCT-derivable foveal pit measures. Second, the UKB predominantly consists of individuals of European ancestries. Consequently, the cohort is not fully representative of global genetic diversity, which may limit discovery opportunities and the generalisability of findings across different populations. Significant correlations in sentinel variant effect estimates, along with associations of EUR-derived polygenic scores in non-EUR cohorts, support a shared polygenic architecture. However, these findings should be interpreted cautiously due to limited sample sizes in the 1KGP-SAS-like and 1KGP-AFR-like groups. To better understand the genetic influences on foveal pit depth, further research in larger and more diverse populations is crucial. Finally, our variant-to-gene approach was designed to be comprehensive, treating all lines of evidence equally. In the context of foveal disease, it may be beneficial to weight certain lines of evidence more heavily. However, the optimal weighting of variant-to-gene criteria is not well established, and the arbitrary assignment of weights could introduce bias.

In conclusion, this study significantly advances our understanding of the genetic basis of human foveal development. Our findings provide a foundation for future functional research into the molecular mechanisms underlying FH and related visual disorders.

## Data Availability

All raw data are available through UK Biobank (https://www.ukbiobank.ac.uk/enable-your-research/apply-for-access).
Summary statistics for the GWAS presented in this paper will be available through GWAS catalog.

## Acknowledgements

We thank all volunteers participating in UK Biobank and who have made this project possible. This study used the ALICE High Performance Computing Facilities at the University of Leicester.

