## Supplementary Figure for "Genome-Wide Insights into the Genes and Pathways Shaping Human Foveal Development"

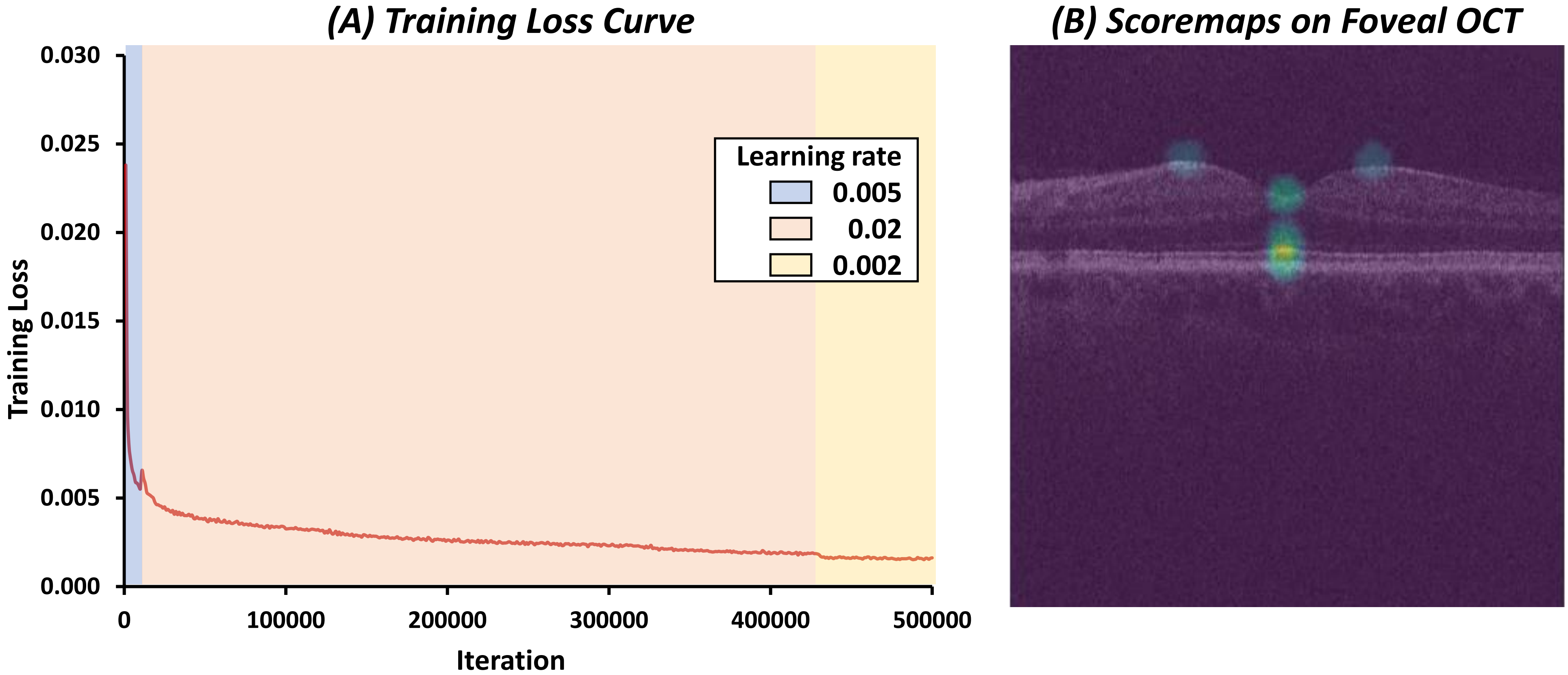


***Supplementary figure 1:*** (A) Training loss curve for custom ResNet-50 model over 500,000 iterations. A staged learning rate schedule was used: 0.005 for the first 10,000 iterations, 0.02 until iteration 430,000, and 0.002 for the final 70,000 iterations. Loss steadily decreased and plateaued at approximately 0.0016, indicating strong convergence. (B) Representative central foveal OCT B-scan overlaid with scoremaps for predicted anatomical landmarks. Warmer colours (e.g., green and yellow) indicate higher model confidence. Shown here are landmark predictions used in foveal depth calculation, including two ILM peak points and the foveal pit. Deeper retinal layers (e.g., IRL, ELM, Ise, RPE) were also predicted.

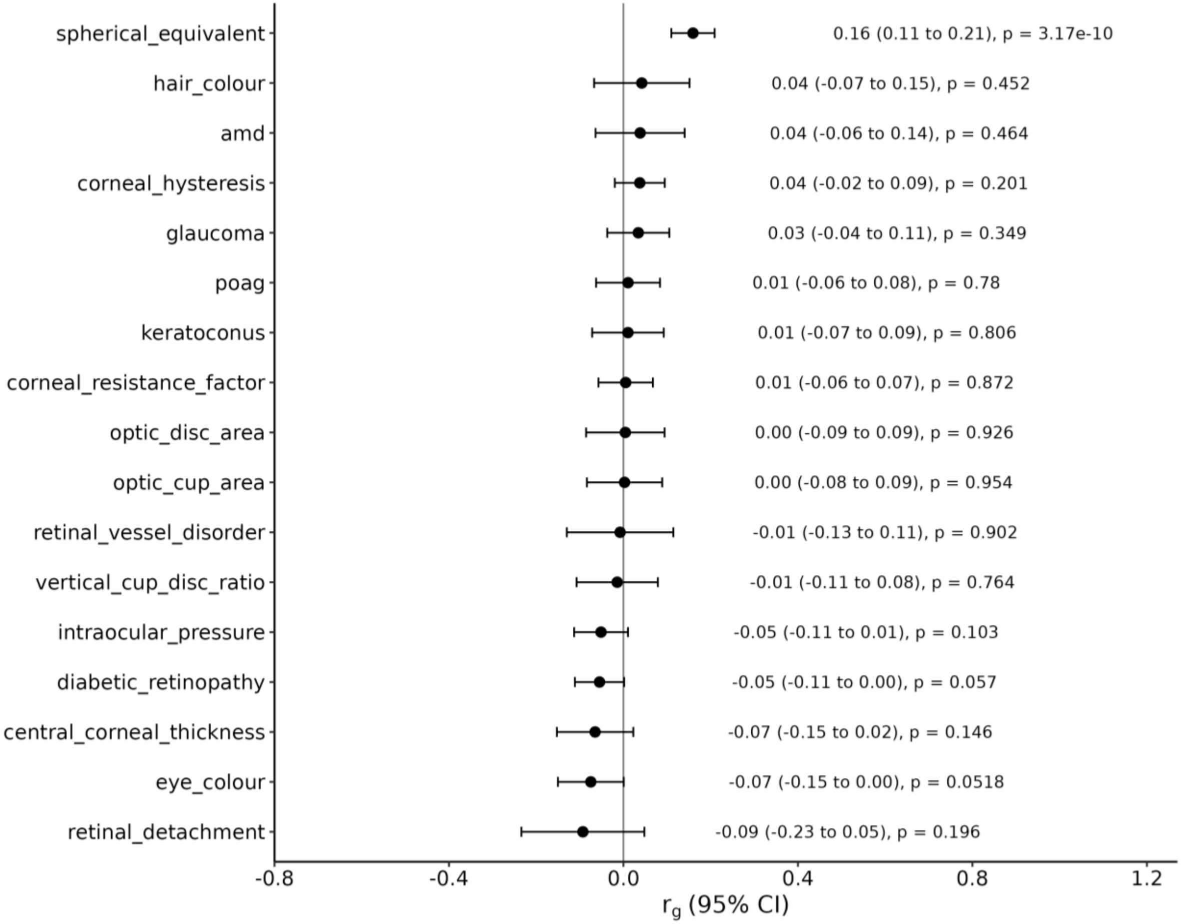


***Supplementary figure 2:*** Genetic correlations (rg) between foveal-pit depth and ocular traits estimated with LDSC. Points denote the estimated *rg and* horizontal bars show the 95% confidence intervals. Plot labels denote the *rg* value, confidence interval and *P-value*
